# AI/ML Models to Aid in the Diagnosis of COVID-19 Illness from Forced Cough Vocalizations: Good Machine Learning Practice and Good Clinical Practices from Concept to Consumer for AI/ML Software Devices

**DOI:** 10.1101/2021.11.13.21266289

**Authors:** K. Kelley, M. Kelley, S. C. Kelley, A.A. Sakara, M.A. Ramirez

## Abstract

From a comprehensive and systematic search of the relevant literature on signal data signature (SDS)-based artificial intelligence/machine learning (AI/ML) systems designed to aid in the diagnosis of COVID-19 illness, we identified the highest quality articles with statistically significant data sets for a head-to-head comparison to our own model in development. Further comparisons were made to the recently released “Good Machine Learning Practice (GMLP) for Medical Device Development: Guiding Principles” and, in conclusions, we proposed supplemental principles aimed at bringing AI/ML technologies in closer alignment GMLP and Good Clinical Practices (GCP).

## 1. Introduction

Artificial intelligence and machine learning (AI/ML) technologies have the potential to augment and democratize health care by providing analysis and insights into the vast amount of health-related data collected pertaining to individuals, as well as their communities. The promise of AI/ML solutions to realize these goals, however, has been hampered by the failure of both Software as Medical Device (SaMD) and Software in Medical Device (SiMD) manufacturers to employ medical device industry standards for design, documentation, testing, manufacturing and deployment across the Total Product Life Cycle (TPLC).

We prospectively planned a PRISMA 2020 systematic review of the relevant literature to demonstrate reproducibility of the published models by building each COVID-19 diagnostic software system for which sufficient details were reported and to conduct head-to-head evaluations both across the completed models and in comparison, to our own AI/ML system. As a secondary objective, our intent was to determine a literature-derived performance goal (PG) for the completed models for comparison to our device with the same intended use.

## 2. Background

Poor data quality leading to poorly trained AI/ML models that cannot perform in a clinical setting has been described extensively in the literature [1-4]. Inadequate sample sizes for training, validation, and testing are often secondary to the lack of availability of datasets that results in a lack of statistical significance and subsequent rejection by informed clinicians.

Furthermore, authors within the current literature have not utilized a prospective plan for the total product life cycle (TPLC) of their device - a necessity for regulatory approval and deployment in the clinical world - promoting a lack of prospective objectives for intended use. The literature also demonstrated a lack of ability of the current models to meet minimum standards set forth by the U.S. Food and Drug Administration (FDA) and the World Health Organization (WHO) for safety and performance. Along with the risks of bias stemming from socioeconomic disparities in data collections, each of these gaps echo statements made in current editorials presenting the challenges of advancing AI/ML technology to aid in the diagnosis of COVID-19 [1] [4].

On 29 October 2021, FDA, CanadaHealth and the Medicines and Healthcare products Regulatory Agency (MHRA) jointly developed and published 10 guiding principles for “Good Machine Learning Practice (GMLP) for Medical Device Development: Guiding Principles,” for the TPLC of SaMDs and SiMDs [5]. The GMLP is a companion to the 13 guiding principles for Good Clinical Practices (GCP) for medical device clinical trials published by the International Conference on Harmonization (ICH) and endorsed by FDA in 1997 [6-7]. With the GCP, standards were set for the design, conduct, and reporting of clinical trials and is key to bringing new medical devices to market. This article will discuss how these principles can be applied to the TPLC to align AI/ML development with clinical trial design and endpoints.

## 3. Materials and Methods

Prior reporting of our methodology and results were detailed in K. Kelley et al. [K. Kelley et. al. 2021 – Manuscript Submitted for Publication] and have been reproduced below in part for ease of reference.

As previously described by our team, we conducted systematic searches of the relevant literature on 12 October 2021 and updated on 7 November 2021, for the purpose of presenting a comparative evaluation of AI/ML systems designed to aid in the diagnosis of COVID-19 from FCV. Searches of the peer-reviewed literature were prioritized but, given the collaborative “shareware” culture of the AI/ML and Data Science communities, pre-print servers were searched for possible contributions. EndNote 2020 was the designated reference manager and PubMed was searched via this software. Serial searches of “Any Field” in PubMed, “Full Text and Metadata” in the IEEE Xplore digital library of the Institute of Electrical and Electronics Engineers, (ieeexplore.ieee.org), “All Fields” in the arXiv open-access archive (arxiv.org), and “Full Text or Abstract or Title” in bioRxiv and medRxiv (medrxiv.org) were performed using the identical search terms as listed below:

- covid and classifier
- covid and neural network
- covid and cough and artificial intelligence
- covid and cough and AI
- covid and cough and machine learning
- covid and cough and ML
- covid and cough and classifier
- cough and neural network
- forced cough vocalization

The results from these serial searches were combined and systematically filtered to achieve a final article pool from which all references would be evaluated for contribution to the stated objectives. Following a basic PRISMA 2020 workflow, our search methodology is illustrated below in Figure 1:

**Figure 1:**
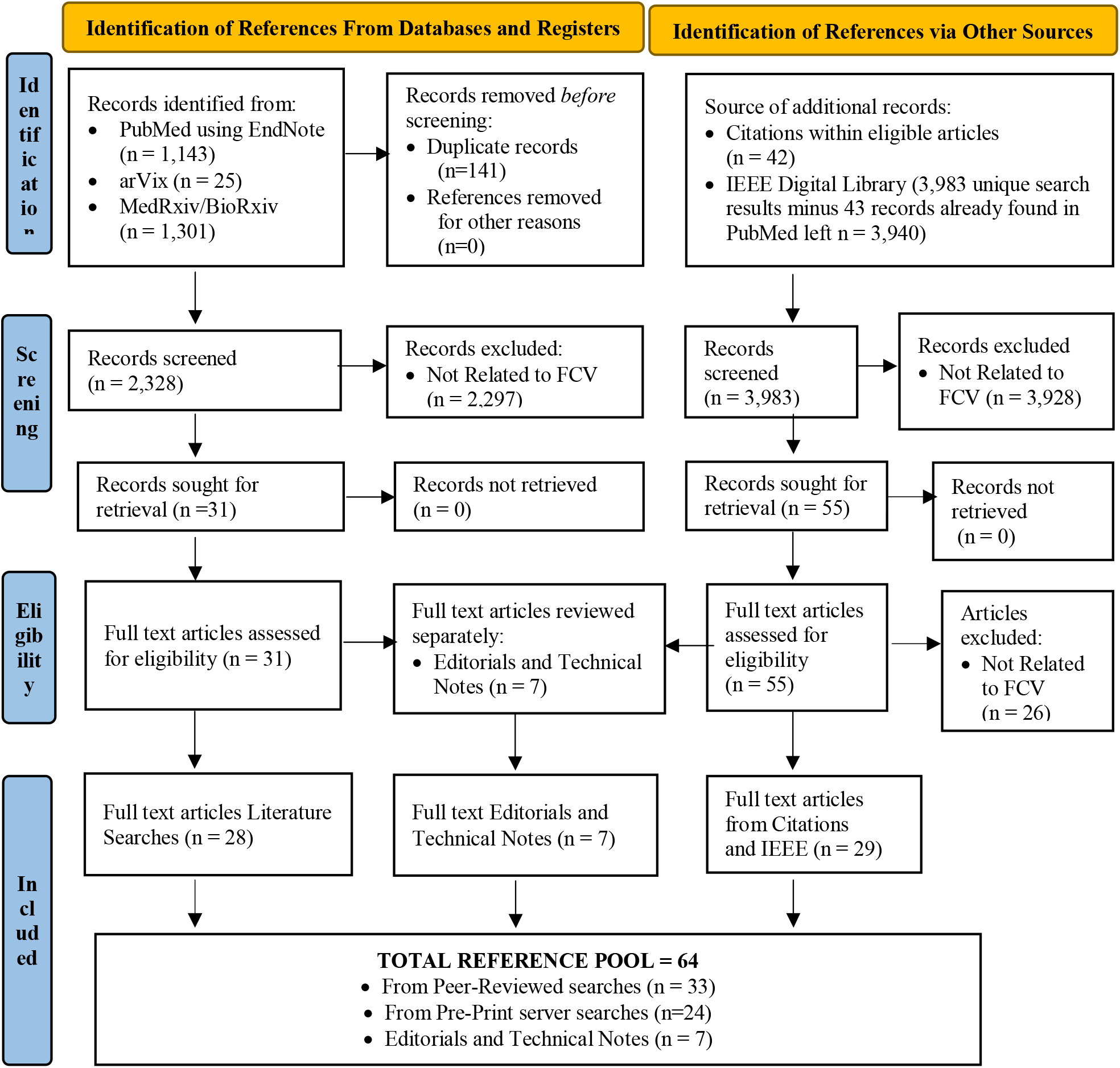
PRISMA 2020 Flow Diagram for New Systematic Reviews (inclusive of Databases, SDS Libraries, and Other Sources)

## 4. Results

From the final pool of 64 references, the remaining 33 peer-reviewed articles and 24 pre-print server records were read and evaluated in full across a multidisciplinary team of Data Scientists, Biotechnical Engineers, Healthcare Clinicians, Product Development and Clinical/Regulatory Affairs professionals. Although each of 57 references purported to include a fully-detailed AI/ML solution, only 14 contained enough information for us to attempt building the stated solution. Of those 14 references, only two peer-reviewed articles demonstrated statistical significance consistent with the requirements for a regulatory pathway.

### 4.1. Primary and Secondary Objectives

No single reference included sufficient details to build a complete model or system, rendering comparisons across models (including our own) unattainable. The primary objectives of the review, therefore, could not be met.

Determination of a literature-derived Performance Goal (PG) was the secondary objective of the original review and is a familiar endpoint in regulatory pathways. None of the references met all inclusion criteria, however, rendering the secondary objective unattainable.

Since the intended objectives of our original systematic review could not be met, we undertook to compare the two peer-reviewed articles [8-9] that demonstrated statistically significance data sets consistent with the requirements for a regulatory pathway and to our model in development. These comparisons will be presented in relationship to two frameworks: the principles of the recently released GMLP and the predicate principles of the GCP for Medical Devices.

### 4.2. GMLP Analysis

The new proposed primary objective was to compare the two articles previously identified as having Level 2a design, Moderate Grade data, and a statistically significant test set size for head-to-head comparison to each other and also to our model in development.

**Table 1:**
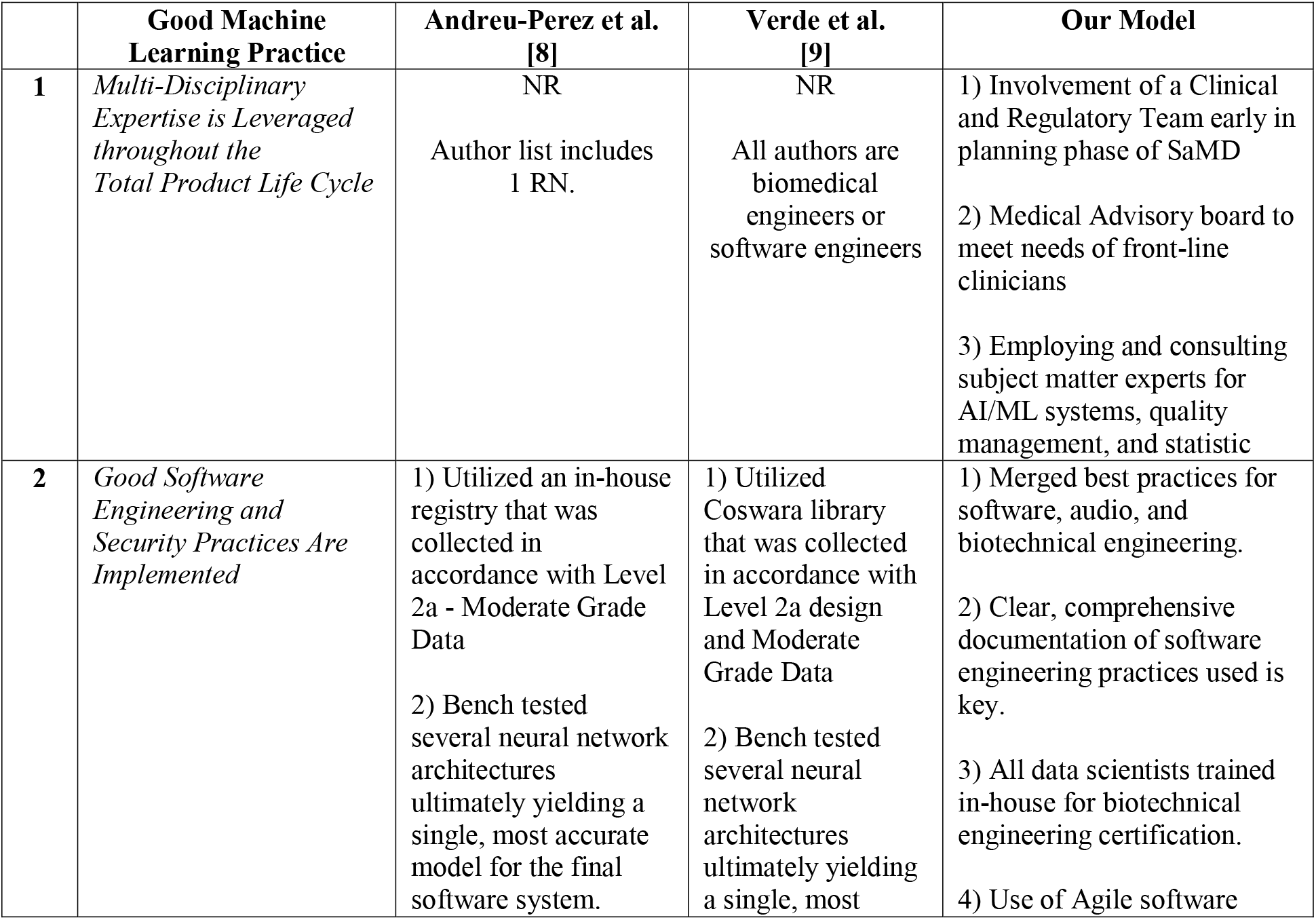
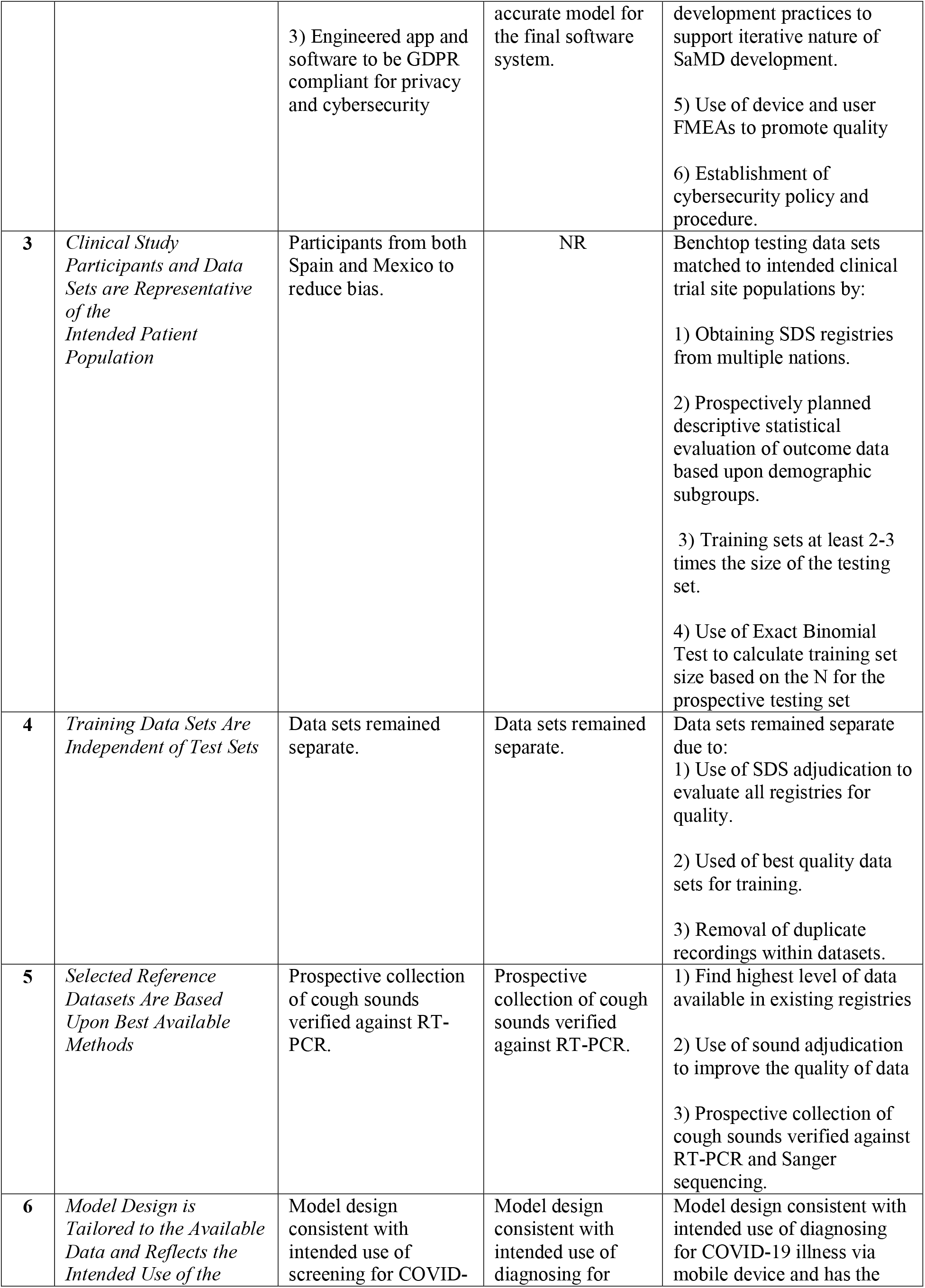
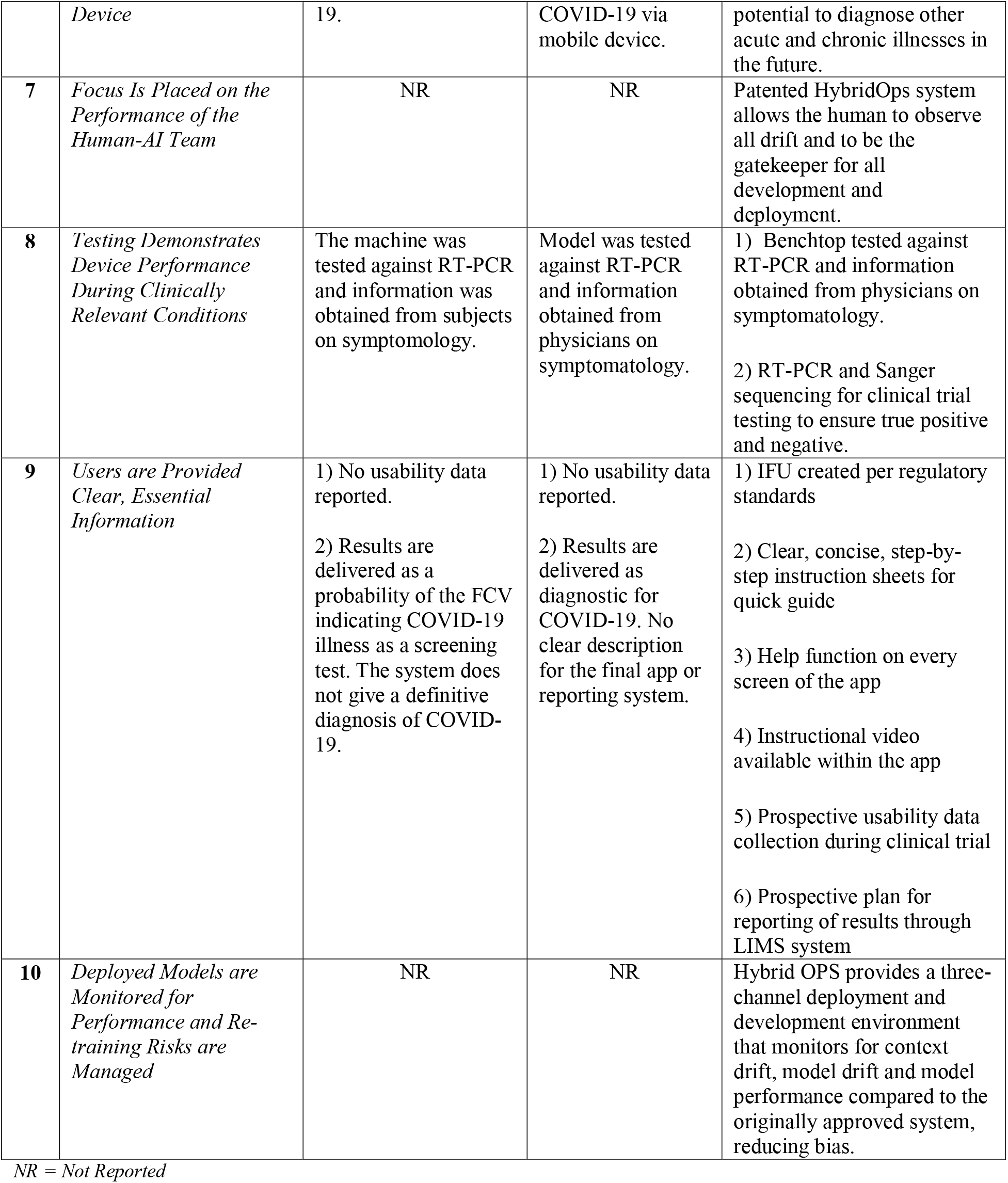
Comparisons to GMLP.

### 4.3. GCP Analysis

Once an AI/ML medical device is developed and ready for clinical testing, there are 13 Good Clinical Practice (GCP) principles that must be observed by international law. AI/ML is not universally accepted in healthcare and problems encountered along the way from inception to deployment of a diagnostic SaMD are varied and many. Changing the level of acceptance will require adherence to Good Clinical Practice. Our next objective was to compare the same two articles head-to-head with our model evaluating GCP.

**Table 2:** Comparisons to GCP.

|  | **Good Clinical Practice** | **Andreu-Perez et al. [8]** | **Verde et al. [9]** | **Our Model** |
| --- | --- | --- | --- | --- |
|  | **Ethics** |  |  |  |
| **1** | *Clinical trials should be conducted in accordance with ethical principles that have their origin in the Declaration of Helsinki, and that are consistent with GCP and the applicable regulatory requirement(s).* | Stated compliance with the Declaration of Helsinki | NR<br>All authors are biomedical engineers or software engineers | Stated compliance with the Declaration of Helsinki |
| **2** | *Before a trial is initiated, foreseeable risks and inconveniences should be weighed against anticipated benefit for the individual trial subject and society. A trial should be initiated and continued only if the anticipated benefits justify the risks.* | Risks NR<br>Benefits well described. | Risks NR<br>Benefits well described | Full QMS documentation to include user and device FMEA complete before trial initiated |
| **3** | *The rights, safety and well-being of the trial subjects are the most important considerations and should prevail over interest of science and society.* | No obvious violation of rights within the article; Not fully reported with no risk analysis. | No obvious violation of rights within the article; Not fully reported with no risk analysis | Full compliance with all international and United States provisions for subject protection, subject rights, minority and underserved access/inclusion and subject safety. |
| **4** | *Nonclinical and clinical information supports the trial.* | Review of literature and related work was completed | Review of literature and related work was completed | Detailed literature reviews undertaken throughout development of model. Benchtop testing to simulate clinical trial performed until all benchmarks for trial met on benchtop. |
|  | **Protocol and Science** |  |  |  |
| **5** | *Clinical trials should be scientifically sound, and described in a clear, detailed* *protocol* | A detailed protocol was written prior to initiating the clinical trial | No description of a clinical trial | A detailed protocol was written prior to initiating the clinical trial |
|  | **Responsibilities** |  |  |  |
| **6** | *A trial should be conducted in compliance with the protocol that has received prior institutional review board (IRB) / independent ethics committee (IEC) approval/favorable opinion.* | The clinical trial was approved by local IECs. | NR | The trial was approved by clinical by a national IRB and accepted by the local IRB. |
| **7** | *The medical care given to, and medical decisions made on behalf of subjects should always be the responsibility of a qualified physician or, when appropriate, of a qualified dentist.* | NR<br>No physician listed in author list. | NR<br>No physician listed in author list. | Clinical trial with physician PIs and Nurse Site Coordinators |
| **8** | *Each individual involved in conducting a trial should be qualified by education, training, and experience to perform his or her respective task(s).* | RNs used to administer testing was appropriate. No other reports of qualifications. | NR<br>All authors are biomedical engineers or software engineers | RNs and trained clinical research assistants overseen by the trial site and responsible for appropriately obtaining the samples.<br>Designated site and sponsor physician PIs for the entire trial.<br>Site Coordinators on site throughout the trial. |
|  | **Informed Consent** |  |  |  |
| **9** | *Freely given informed consent should be obtained from every subject prior to clinical trial participation.* | Full informed consent obtained on all participants. | NR | Full informed consent obtained on all participants. |
|  | **Data Quality and Integrity** |  |  |  |
| **10** | *All clinical trial information should be recorded, handled, and stored in a way that allows its accurate reporting, interpretation and verification* | GDPR compliant data collection and storage | NR | All data collected and maintained in HIPPA compliant fashion, All results reported as HL7 compliant reports in HIPPA compliant fashion. System designed to meet all medical device and SaMD ISOs, standards, good practices, and cybersecurity benchmarks |
| **11** | *The confidentiality of records that could* | Researchers ensured patient confidentiality | NR | Researchers ensured patient confidentiality during the trial. |
|  | *identify subjects should be protected, respecting the privacy and confidentiality rules in accordance with the applicable regulatory requirement(s).* | during the trial. GDPR compliant app developers designed to this end. |  | HIPPA/HL7 compliant app and API developers designed to this end. |
|  | **Investigational Products** |  |  |  |
| **12** | *Investigational products should be manufactured, handled and stored in accordance with applicable Good Manufacturing Practice (GMP). They should be used in accordance with the approved protocol.* | See Table #1 (GMLP) | See Table #1 (GMLP) | See Table #1 (GMLP) |
|  | **Quality Control / Quality Assurance** |  |  |  |
| **13** | *Systems with procedures to ensure quality of every aspect of the trial should be implemented* | NR | NR | Full QMS system in place and documentation complete before trial begin |

NR = Not Reported

## 5. Discussion

The publication of GMLP that has followed the GCP for Medical Devices raises the bar for the scientific rigor of SiMD/SaMD development, the associated benchtop testing, and clinical trials. Collectively, these principles reinforce that prospective design applies not only to the design of the software but also the testing and, ultimately, the evidence-based clinical trial of the device.

Our review of 57 articles and reports describing FCV-SDS based COVID-19 diagnostic software components and systems found that only 2 of the 57 articles could meet Level of Evidence, grade of data and statistical significance of testing criteria. When these 2 articles were evaluated against the principles for GMLP and GCP, only the article by Andreu-Perez et al. demonstrated a degree of compliance with the guidelines [8].

Our own experience, albeit pre-clinical trial, found that the GMLP and GCP guidelines are well aligned with good engineering practices and good evidence-based medicine research practices. We respectfully submit the following supplements to the GMLP and GCP to include preclinical benchtop testing principles, techniques to ensure AI/ML model consistency and guidance for overcoming data borders:

- Prior to Clinical Trial, investigational AI/ML software devices should be trained and validated using a reference dataset that is clinically relevant to the proposed clinical trial. Furthermore, the model should demonstrate successful benchtop testing using a testing dataset that is clinically relevant and with testing endpoints that are identical to prospectively designed clinical trial. This will promote adherence to two of the current GMLP principles - separation of training and test sets and providing for the highest quality of data.
- The trained and validated AI/ML models used in pre-clinical trial benchtop testing and for clinical trials should be reproducible by independent researchers. This requires that the source code for the complete model, including post training transfer learning, is made available as part of peer-reviewed published reports to facilitate replication of the benchtop testing and/or clinical trials. The use of original source code and transfer learning ensures that the replicated tests and trials are using the identical AI/ML model, making direct comparisons possible.
- Clinical trials that cross data borders and parallel AI/ML models operating in different data boundary regions should implement the same source code and transfer learning in all data regions. Additionally, if the clinical trial crosses data borders, parallel AI/ML models operating in different data boundary regions should synchronize transfer learning using federated data and swarm learning. This approach would respect applicable data boundaries, which is essential for international operations. Without these considerations, data borders will impose healthcare inequities and limit access.

## 6. Conclusion

Adherence to GMLP and GCP by AI/ML SaMD and AI/ML SiMD developers will also serve to improve the confidence of healthcare providers and healthcare decision makers in these new and promising technologies. The newly released GMLP guidelines provide an ideal framework for AI/ML and SiMD/SaMD developers to guide their efforts throughout the TPLC, yielding more meaningful clinical outcomes. Data scientists, clinicians, and regulatory specialists must collaborate to ensure that good practices translate into a product that meets the needs of both healthcare providers and patients. Focusing on quality and incorporating regulatory guidance throughout the TPLC will allow AI/ML developers to bring a high-quality medical to market.

GCP principles provide the guidance for clinical trials to be conducted ethically and scientifically with clear and detailed protocols. Care must be given by appropriately qualified personnel with adequate experience. Most importantly, documentation of adherence to these practices is key, with records easily accessible and retrievable for accurate reporting, verification and interpretation.

In order to bring new AI/ML technology solutions to healthcare, developers must prospectively plan to use the GMLP and GCP principles from the beginning of the TPLC.

## Data Availability

All data produced in the present work are contained in the manuscript

## Notes

### Competing Interest Statement

K. Kelley, MD, FAADM, Chief Regulatory Medical Director for RAIsonance, Inc.,
M. Kelley, MSN, RN, LNC, Co-Chief for Clinical and Regulatory Affairs for RAIsonance, Inc.,
S. C. Kelley, DNP, AGACNP-BC, FNP-BC, SVP Clinical Trials and Quality Improvement for RAIsonance, Inc.
A. A. Sakara, NP, MSN, RN, PHRN, Co-Chief for Clinical and Regulatory Affairs for RAIsonance, Inc.,
M. A, Ramirez, DO, PhD, Chief Medical Officer, Chief Scientist, Chief Strategy and Planning Officer for RAIsonance, Inc

### Funding Statement

RAIsonance, Inc.

